# Dynamics of Vaccine-Hesitant Parents’ Considerations Regarding Covid-19 Vaccination

**DOI:** 10.1101/2022.10.02.22280627

**Authors:** Erga Atad, Itamar Netzer, Orr Peleg, Keren Landsman, Keren Dalyot, Shanny Edan Reuven, Eyal Nitzany, Ayelet Baram-Tsabari

## Abstract

**Introduction:** Most studies present a snapshot of hesitant parents’ decisions and thinking concerning COVID-19 vaccination, but for many it is a dynamic rather than a stable process. We examined the considerations of a group of vaccine hesitant parents (VHPs) with respect to COVID-19 vaccinations for their children before, during and after the main vaccination campaign for the 12 to15-year-old age group in Israel, over a six-month period.

**Methods:** Digital surveys were administered to 1118 Israeli parents. After VHPs were identified, three surveys were conducted to evaluate considerations that discourage or encourage vaccination. A logistic regression was carried out on sixteen models; of these, six were found to be statistically significant.

**Results:** 456 parents’ data were analyzed. Parents’ intentions to vaccinate prior to the vaccination campaign were a good predictor of their actual behavior, (rp=.497, p<.001). We divided the parents into four groups: consistently pro-vaccine (39.4%), consistently anti-vaccine (15.2%), pro-vaccine parents who did not vaccinate (17.6%), and anti-vaccine parents who did vaccinate (27.9%). We identified eight considerations that were significant in VHPs’ vaccination behavior: trust in scientists, doctors and drug companies, children’s preferences, spread of COVID, social responsibility, children’s characteristics, the vaccine’s speed of development and its side effects.

**Discussion:** Greater vaccine uptake for teenagers may depend on the attitudes and perceptions of their parents. We identified encouraging and discouraging considerations that may make potential targets for public health officials when communicating about vaccines.

**Article Summary:** Encouraging and discouraging considerations re COVID-19 vaccination are explored and identified in vaccine hesitant parents.

**What’s known on this subject:** Most studies found that vaccine uptake is improved among well-to-do or highly educated parents. Severity of the disease and safety of the vaccination are the main factors influencing the decision.

**What this study adds:** The dynamics of attitude change toward vaccination among VHPs may be affected by several different factors. Chief are children’s preferences and individual health characteristics, side effects and speed of vaccine development

## Introduction

Israel has led a rapid vaccine rollout campaign to combat COVID-19, starting with vaccination of medical personnel, high risk populations and senior citizens^1^. This has resulted in close to six million people receiving both Pfizer-Biontech COVID-19 vaccination doses, out of a total population of 9.217 million (5,783,404, 62.74%^2^.) As of early December 2021, the third dose, or booster, had already been administered to 4,104,491 individuals (44.52%). The success of the vaccination campaign for 12–15-year-olds, who make up 6.7% of the population^3^, rests on communicating the necessity, safety, and efficacy of the vaccine to them and their legal guardians. As of December 2021, the rates of full vaccination (two shots up to six months apart, or two shots and a booster six months later), were 58.47% (0.01% with the booster shot) in the 12-15-year-old age group, and 64.40% (14.71% with the booster shot) in the 16–19-year-old age group. The rate of vaccination at the beginning of the 12-15 vaccination phase was challenged by the relative remission of the epidemic, the rollback of most public health measures in Israel at that time, the low morbidity associated with COVID-19 infection in children^4^, and the rapid rollout of a new vaccine under FDA Emergency Use Authorization (EUA) rather than full FDA approval^5^.

In order to study the dynamics of parents’ considerations, we examined a group of vaccine hesitant parents (VHPs) regarding COVID-19 vaccinations for their children at three time points: before, during and after the main vaccination campaign for the 12–15-year-old age group following the third COVID-19 wave (most patients at this stage were infected with the Alpha variant, but Delta was already beginning to spread). The ongoing results were communicated to Israeli policymakers and public health officials in real time to contribute to informing their decisions regarding the vaccine rollout and public communication, through Mida’at, a non-government organization and member of the Public COVID-19 Advisory Board to the Israel Ministry of Health.

Vaccine hesitancy was defined by the Strategic Advisory Group of Experts (SAGE) Working Group on Vaccine Hesitancy as a delay in acceptance or refusal of vaccination despite the availability of vaccination services ^6^. Several factors have been found to contribute to hesitancy including misleading information, beliefs and perceptions about vaccines and negative attitudes and behavior toward vaccination, alongside demographic characteristics (large families or older children, low levels of income and education)^7,8^. VHPs are also concerned about vaccine safety, are more likely to believe that children receive too many vaccines, that their child may have serious side effects from a vaccine, and that vaccines can negatively impact a child’s immune system.^9^

Studies focused on COVID-19 vaccination indicate more focused fears and considerations related to vaccine hesitancy. A pre-approval study found that guardians wishing to vaccinate their children cited protecting them against COVID-19 as the main reason, while those refusing future vaccination reported the vaccine’s novelty as being their main reason for refusal^10^. A trial involving the responses of 1,313 individuals found that the characteristics of the “maybe” respondents (i.e., ‘maybe I’ll get vaccinated’) differed from those of the “yes” or “no” respondents. Respondents were more likely to respond “maybe” than “yes” if they thought COVID-19 was less severe, distrusted science, or were less willing to vaccinate for Influenza^11^. Additional studies have found correlations between willingness to vaccinate oneself against COVID-19 and the perceived risk of the disease, or perceived safety of the vaccine, respectively^12^. Socioeconomic conditions were also found to be in correlation with vaccination behavior^13,14^.

Unlike most studies that present a snapshot of parents’ decisions and thinking regarding COVID-19 vaccination, for many it is a dynamic rather than a stable process. A moving target of concerns informed by daily news against the backdrop of changing health threats.

The present study followed parents at three time points, to profile those who changed their mind. We aimed to identify the reasons positively and negatively affecting the decisions of parents who were indecisive or hesitant about their decision to vaccinate, to contribute to better informing public health officials and healthcare workers when communicating the importance of vaccines to VHPs.

## Methods

Institutional review board approval was obtained from the Technion - Israel Institute of Technology (2021-034; 2021-056; 2021-089). We conducted a three-stage study which included data collection using digital surveys via a commercial online data collection provider (iPanel, Bnei Brak, Israel), as shown in Figure 1. A link was provided to a Qualtrics survey. Total compensation for each survey did not exceed 20 NIS (about 6 US dollars). Informed consent was digitally obtained at the beginning of each questionnaire.

**Figure 1:**
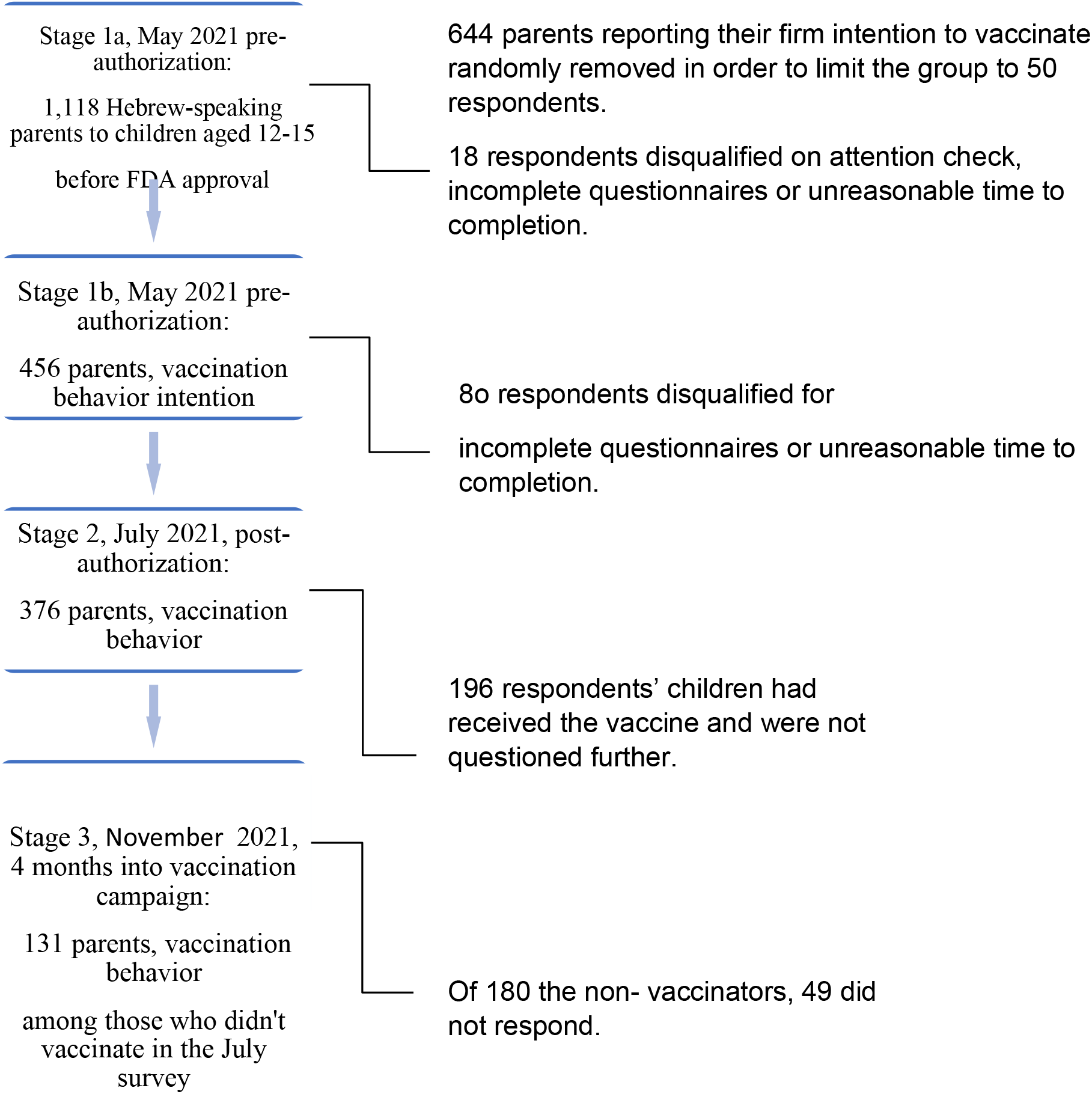
Study design: four stages of the study.

**Figure 2.**
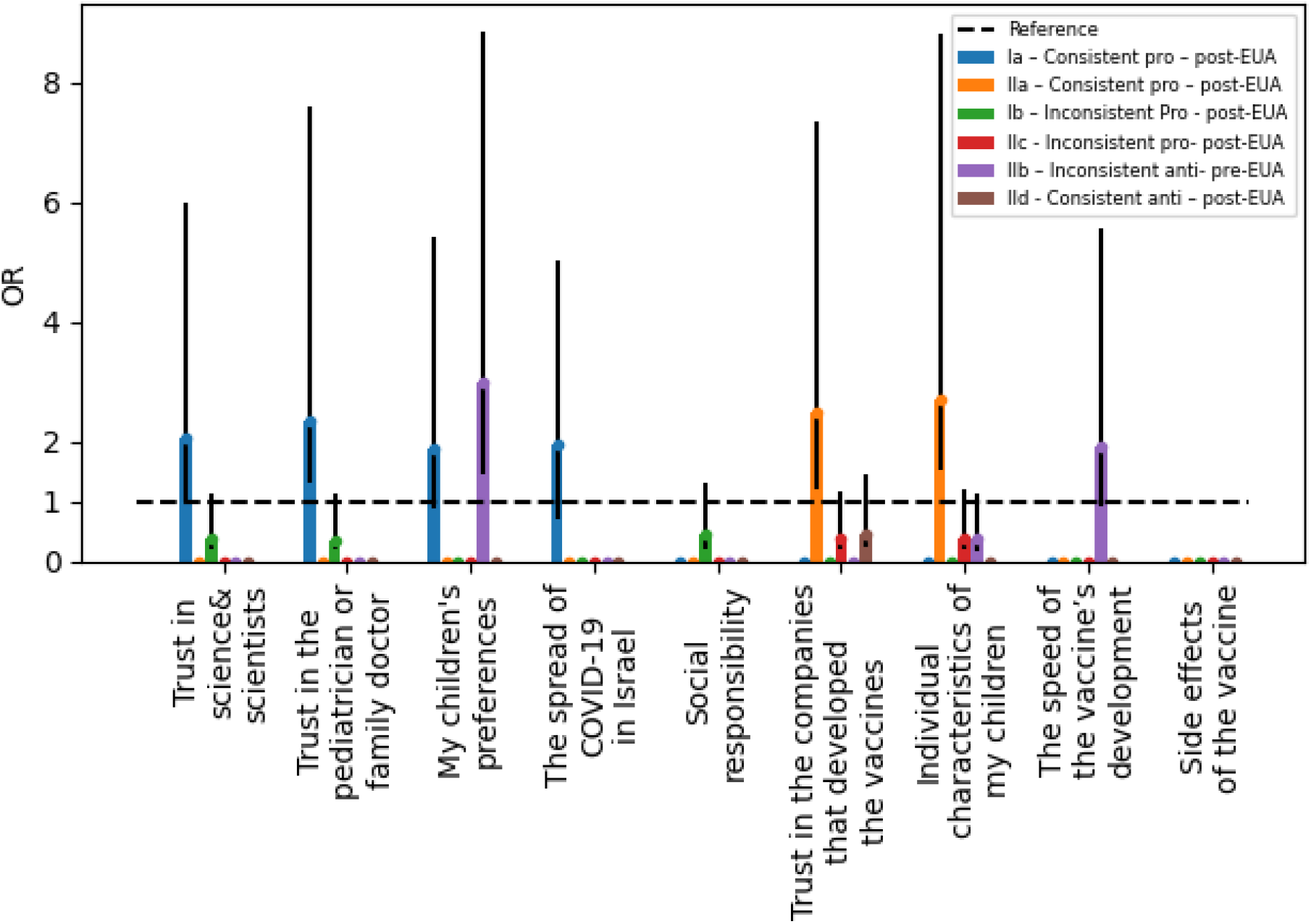
Odds Rations of Considerations by Models Odds ratio of significant considerations that associate people to a certain group. Only significant considerations (x axis) appear, mean values (odds ratio; y axis) are presented with their error bars. Over the threshold of 1 (dashed line) - the consideration characterizes the group, under the threshold - the consideration does not characterize the group. This means that it is more likely that people that belong to this group did not mark it. For example, people who belong to the group Consistent pro vaccination (model Ia; marked in blue) are more likely to trust science and scientists - blue bar on the left is above the reference threshold. However, people who belong to the group Inconsistent: Pro-vaccination who did not vaccinate (model IIa; marked in orange) are less likely to mark this argument - orange bar on the left is below the reference threshold.

This study was based on an initial survey that took place from April 28 to May 6, 2021, prior to the FDA approval of vaccination for 12-15-year-olds on May 10th, 2021^5^, given that the vaccination campaign in Israel was expected to commence shortly afterwards. The second stage of the survey was administered on July 13 to July 21, 2021, several weeks after the vaccine was approved in Israel (June 2, 2021), and the third in November to allow for late adopters to reach a decision.

### Respondent accretion

Exclusion of respondents throughout the study is presented in Figure 1. Of the 1118 parents who responded to stage 1a (below) 644 indicated they would definitely vaccinate their children. A small number of parents who had definite intentions for or against vaccination (scored 1 or 5 on the Likert scale in stage 1a) were left in the analysis for the sake of comparison. We limited the “definitely will vaccinate” group included in the second stage to 50 randomly chosen respondents. We had no need to limit the “definitely will not vaccinate” group as it numbered fewer than 50 (6.3% in stage 1a). While the original cohort was chosen to represent the demographics of Hebrew speaking Israelis (for the sake of linguistic simplicity) the resulting sample was relatively more secular and more highly educated compared to the entire Hebrew-speaking population (Table 1).

**Table 1:** Demographic Characteristics of the Survey Population.

| **Variables** | **Categories** | **Pre/post EUA**<br>**Sample (%)** | **4 months into**<br>**campaign**<br>**Sample (%)** |
| --- | --- | --- | --- |
| Gender<sup>1</sup> | Women | 51 | 50.4 |
|  | Men | 49 | 49.6 |
| Religiosity | Secular | 57.2 | 48.1 |
|  | Traditional | 20.5 | 21.4 |
|  | Religious | 16.2 | 21.4 |
|  | Ultra-Orthodox | 6.1 | 9.2 |
| Highest level<br>of education | No high school matriculation<br>certificate | 12.2 | 16.8 |
|  | High school matriculation<br>certificate | 20 | 20.6 |
|  | Post-secondary program,<br>without academic degree | 17.3 | 16.8 |
|  | Academic degree | 50.5 | 45.8 |
| Income<sup>s</sup> | Below average | 24.2 | 29.9 |
|  | Average | 34.3 | 39.4 |
|  | Above average | 39.1 | 30.7 |
| Total |  | 376 | 131 |

### Research Tools

The digital surveys were developed by the research team based on earlier COVID-19 related surveys^8,15,16^ and the recommendations of the SAGE Working Group on Vaccine Hesitancy^6^. The content validity of the research tools was determined by expert professional judgment (feedback from ten experts in public health and in science communication).

Stage 1 - (before FDA EUA) included a single question that examined intention to vaccinate. Respondents were asked to answer a 5-point Likert question: “Do you intend to vaccinate your children when the COVID-19 vaccine becomes available for them?”. A sample of 1118 Hebrew-speaking Israelis who are parents of 12-15-year-olds consented to complete this stage. Parents were chosen by iPanel from their representative sample, from all areas of Israel.

Following the previous question, we asked 456 respondents to specify the most influential considerations in their intention to vaccinate or not to vaccinate their children. They were asked to choose their five most significant considerations in favor of vaccination, and their five most significant considerations against vaccination from a list. The survey also included questions related to additional information such as gender, education level, income level, and religiosity.

In stage 2 (After FDA EUA and during the vaccination campaign) we returned to our sample of 456 from stage 1b to examine their actual vaccination behavior. Of these, 376 completed the questionnaire in no less than 3 minutes. The remaining 80 parents were disqualified (see Figure 1). The respondents who reported that they had vaccinated their children were asked to specify the considerations that convinced them to vaccinate, and their main considerations against vaccination. Participants who reported that their children were not vaccinated, were asked to specify the considerations that convinced them not to vaccinate, and the most significant considerations in favor of vaccination. We also attempted to identify parents who changed their minds between study stages (respondents who intended to vaccinate but did not, and respondents who intended to refrain from vaccination but did vaccinate their children). Of these 456 parents, 180 (39.5%) reported that they did not vaccinate.

We returned to these 180 parents in the 3rd stage (4 months after commencement of the vaccination campaign) in order to isolate late responders and further inquire whether they had vaccinated their children and their considerations had been. A total of 131 parents responded to the final survey. No information was collected on why 49 parents declined to respond. We surmise that “survey fatigue” was involved.

### Stratification of parent groups

To differentiate between parents who changed their mind and those who were consistent, we split the sample into four parental groups according to the coherence between their initial intention in May and actual behavior later in 2021 (Table 3):

I. Consistent pro-vaccination - those who reported their intention to vaccinate and did so in practice.
II. Inconsistent: anti-vaccination who did vaccinate - those parents who reported they did not intend to vaccinate their children or were hesitant but did vaccinate in practice.
III. Inconsistent: Pro-vaccination who did not vaccinate - those parents who reported they intended to vaccinate their children or were hesitant but did not do so in practice
IV. Consistent anti-vaccination - those who reported their intention not to vaccinate and did not vaccinate in practice.

We examined each group separately to differentiate what encouraged or discouraged them to vaccinate.

### Statistical methods

Sixteen binary logistic regressions were run:

A. Four analyzed the Stage 1 survey (before FDA EUA) and another four analyzed the Stage 2 survey (during the vaccination campaign post FDA EUA), to determine whether a set of 20 considerations rated as encouraging vaccination were associated with the likelihood of each respondent to be in one of the four parental groups.
B. Four regression models analyzed the Stage 1 survey (before FDA EUA) and four analyzed the Stage 2 survey (during the vaccination campaign post FDA EUA), to examine whether a set of 20 considerations rated as discouraging vaccination were associated with the likelihood of each of respondent to be in one of the four parental groups.

## Results

Overall, 253 (67.3%) of the 376 parents surveyed chose to vaccinate their teenagers. Of these, 196 did so during the main vaccination campaign and another 57 did so by the end of it. Parents’ intentions to vaccinate before the vaccine’s authorization was a very good predictor of their actual behavior later that year, (rp=.497, p<.001). Table 2 depicts the relationship between parents’ intentions and actual self-reported vaccination behavior.

**Table 2:** Parents’ vaccination intentions in May 2021 vs. vaccination behavior in July and November 2021.

|  | May- (pre-EUA)<br>Intention | July (post-EUA) - Behavior |  |  | November (4 months into campaign) - Behavior |  |
| --- | --- | --- | --- | --- | --- | --- |
| Behavior in July & November<br>Intention in May | May & July total | Vaccinated | Did not vaccinate | November total | Vaccinated | Did not vaccinate |
| Definitely will vaccinate | 39 | 33 (84.6%) | 6 (15.4%) | 5 | (80%) 4 | (20%) 1 |
| Probably will vaccinate | 134 | 93 (69.4%) | 41 (30.6%) | 26 | 18 (69.2%) | 8 (30.76%) |
| I haven't decided yet | 131 | 59 (45%) | 72 (55%) | 55 | 31 (56.3%) | 24 (43.6%) |
| Probably will not vaccinate | 43 | 9 (20.9%) | 34 (79%) | 28 | 3 (10.7%) | 25 (89.2%) |
| Definitely will not vaccinate | 29 | 2 (7%) | 27 (93.1%) | 17 | 1 (5.88%) | 16 (94.1%) |
| **Total** | **376** | **196 (52.12%)** | **180 (47.87%)** | **131** | **57 (43.51%)** | **74 (56.48%)** |

**Table 3:**
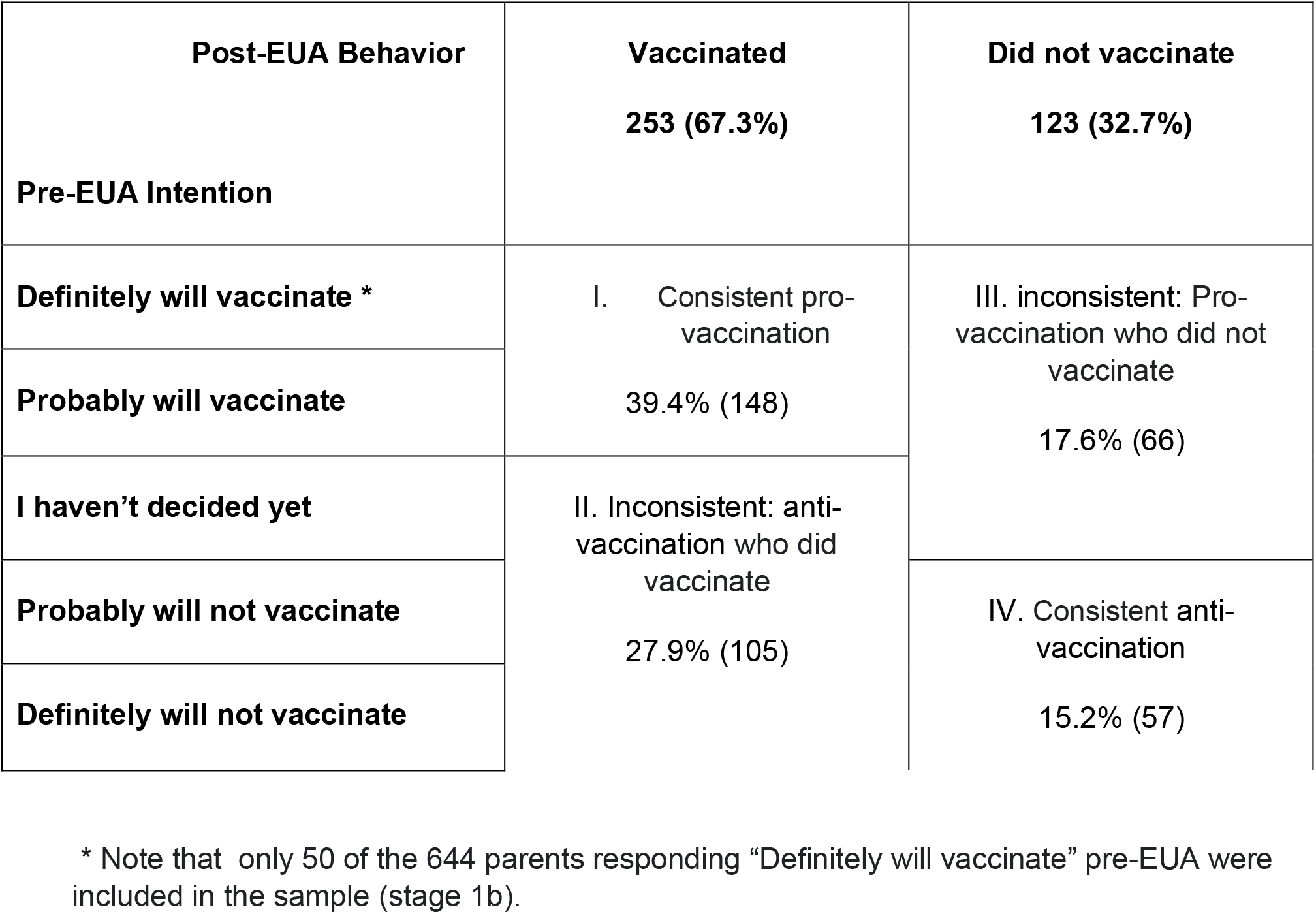
Four parental groups in terms of coherence between their initial intention and actual behavior.

For the sake of brevity, we will only discuss the six regression models that were statistically significant out of the sixteen models. These results are presented in table 4.

### Considerations encouraging vaccination

#### Model Ia - Consistent pro vaccination – Second Survey, post authorization

*Trust in science & scientists* (b=0.72, OR 2.05, p<0.05); *Trust in the pediatrician or family doctor* (b=.85, OR 2.35, p <.05); *My children’s preferences* (b=0.64, OR 1.89, p<.005); and *The spread of COVID* (b=0.67, OR 1.95, p<.005) were all positively statistically significant. This indicates these considerations encouraged parents to be consistent in their decision to vaccinate

#### Model Ib - Inconsistent: Pro-vaccination who did not vaccinate - Second Survey, post authorization

*Trust in science & scientists* (b=-0.97, OR 0.38, p<0.05); *Trust in the pediatrician or family doctor* (b=-1.09, OR 0.34, p<0.05); and *Social responsibility* (b=-0.75, OR 0.47, p<0.05) were negatively statistically significant. VHPs who intended to vaccinate but did not do so, had decreased odds ratios for viewing these considerations as encouraging. This would suggest that non-vaccination correlated with low trust.

### Considerations discouraging Vaccination

#### Model IIa-Consistent pro vaccination – Second Survey, post authorization

*Trust in the companies that developed the vaccines* (b=0.91, OR 2.49, p<0.05), and *Individual health characteristics of my children* (b=.98, OR 2.689, p<0.05) were both positively statistically significant, indicating that the probability of respondents who were intending to vaccinate, to ultimately do so, was higher if considering both as possibly discouraging considerations.

#### Model IIb - Inconsistent anti vaccination who did vaccinate-First Survey, pre-authorization

*Speed of vaccine development* (b=-.92, OR 0.39, p <.05) was negatively statistically significant, whereas *Side effects of the vaccine* (b=.64, OR 1.91, p <.05) and My children’s preferences (b=1.09, OR 2.98, p <.05) were positively statistically significant as potentially discouraging considerations. This indicates that parents who intended not to vaccinate but did so, were not discouraged by the speed of vaccine development, but were discouraged by the vaccine’s perceived side effects, and by their children’s vaccine preferences (despite ultimately vaccinating).

#### Model IIc - Inconsistent pro vaccination who did not vaccinate - Second Survey, post authorization

Trust in the companies that developed the vaccines (b=-.95, OR 0.38, p<0.05) and Individual health characteristics of my children (b=-0.96, OR 0.38, p<0.05) were both negatively statistically significant, indicating that distrust in companies and perceived health characteristics of their children discouraged parents who would have liked to vaccinate, from doing so.

#### Model IId- Consistent anti vaccination – Second Survey, post authorization

*Trust in science and scientists* (b=-1.19, OR 0.30, p<.005), and *Trust in the companies that developed the vaccines* (b=-0.75, OR 0.47, p<.001) were negatively statistically significant. This indicates that distrust of scientists and companies contributed to these parents’ decision not to vaccinate.

## Discussion

Out of 20 considerations rated for their value in encouraging or discouraging vaccine hesitant parents in their decision to vaccinate their teenage children, a small number were found to be statistically significant. Considerations encouraging vaccination:

*Trust in Science and Scientists, Trust in the Pediatrician or Family Doctor, Children’s Preferences* and *the Spread of the Disease* were instrumental in encouraging hesitant parents to vaccinate. *Social Responsibility* was also found to be an encouraging consideration for some parents.

Considerations discouraging vaccination:

VHPs viewed trust (or rather distrust) in the companies developing COVID-19 vaccines, and perceptions of their children as having unique (individual) health characteristics as potentially dissuading considerations. The vaccine’s perceived *Rapid Development* and perceptions of *Unknown Side Effects* were found to discourage VHPs from vaccinating their children, whereas Children’s *preferences* may have increased the likelihood of anti-vaccination-leaning VHPs to go against their initial preference and ultimately vaccinate.

Previous studies have identified the main considerations affecting vaccine uptake or refusal, and Charles Shey et al (2021)^17^ hypothesize that because people’s vaccination views usually “comprise an ongoing engagement that is contingent on unfolding personal and social circumstances”, it can potentially change over time. Our findings indeed show the significance of such a potential change with 45% of our vaccine hesitant parents sample changing their mind. This is a much more substantial group than reported in previous study with mothers at baby-birth (9.7% hesitancy) surveyed again at 24 month (5.9% hesitancy) that found that hesitancy is a dynamic measure peaking around childbirth and may be reduced with experience with vaccines. Our findings present vaccine hesitancy as not static but rather a dynamic condition, with some parents’ considerations changing and resulting in a behavioral change. These may be affected by several different individual and contextual factors, sometimes acting in opposing directions.

## Limitations of this study

We could not identify specific encouraging considerations that characterized parents who belonged to group IV (consistently anti-vaccination). This is partly because many parents who were firmly and consistently anti-vaccination did not check any considerations that could encourage them to vaccinate.

Despite the under-representation of consistent pro-vaccine parents due to limiting the number of parents who reported the intention to “definitely vaccinate” pre-authorization to only 50, the final self-reported rate of vaccination was 67%, whereas the actual vaccination rate of 12– 15-year-olds in Israel in 4 months after commencement of the vaccination campaign was around 62%^18^. This stems from the absence of non-Hebrew speakers in the sample, and an underrepresentation of ultra-orthodox parents. In addition, parents’ perceptions of their children’s preferences was found to be a factor affecting parents’ decisions, but this is limited by the fact that we did not correlate this with the children’s actual preferences.

## Conclusion

Several perceptions or considerations regarding the COVID-19 vaccine and its implications for the vaccination of teenagers were found to significantly affect the decisions and behavior of vaccine-hesitant parents. These may inform the strategies and policies utilized by authorities to increase vaccine uptake. The findings suggest that trust in family physicians and pediatricians may improve vaccine communication if messages are expressed by these health professionals (i.e., as opposed to non-physicians). The teens’ own wishes regarding vaccination also played a role in their guardians’ decisions, thus potentially making it worthwhile to communicate information about the epidemic and vaccine through routes that are accessible to children and teens. Messages regarding the vaccines should directly address parents’ concerns (such as the speed of vaccine development and hypothetical future side effects, as these are considered irrelevant by many immunologists). Finally, initial attitudes and intentions of VHPs do not necessarily represent their actual vaccine behavior, and specific considerations may correlate with changes in this behavior. This may indicate that the reinforcement of encouraging considerations and the rebuttal of discouraging ones may enhance vaccine uptake.

## Data Availability

All data produced in the present study are available upon reasonable request to the authors

## Abbreviations

VHPs: vaccine hesitant parents
FDA: Food and Drug Administration

